# Primary care Screening Questionnaire for Depression (PQS4D)’s dimensionality, internal consistency, nomological network, and gender differential item functioning

**DOI:** 10.1101/2025.03.02.25323200

**Authors:** Adalberto Campo-Arias, John Carlos Pedrozo-Pupo, Carmen Cecilia Caballero-Domínguez

## Abstract

The Primary care Screening Questionnaire for Depression (PQS4D) was recently introduced. It is a four-item instrument based on the International Classification of Diseases (ICD-10). However, psychometric indicators such as dimensionality, nomological validity, and gender differential item functioning are unknown.

The study aimed to test the dimensionality, internal consistency, nomological network, and gender differential item functioning (Gender-DIF) among elderly Colombian COPD patients.

A psychometric study was designed in which 349 COPD outpatients were accepted to participate. They were aged between 60-98 years (M=75.56, SD=8.41); 61.89% were male, and 80.23% presented low deterioration (A or B COPD combined evaluation). The PQS4D (Spanish version), Brief Zung’s Self-rating Depression Scale (BSDS), Brief Zung’s Self-rating Anxiety Scale (BSAS), COPD Assessment Test (CAT), and Frail Non-Disabled [FiND] Screening Tool were completed. Dimensionality (exploratory and confirmatory factor analysis, internal consistency (Kuder-Richardson coefficient and McDonald’s omega), nomological network (Student’s test and Spearman correlation [*r*_*s*_]), and Gender-DIF (Kendall’s tau b correlation [t]) were done.

The PQS4D presented a one-dimension structure that accounted for 55.78% of total variance with adequate fit-of-goodness indicator [Normalized chi-square of 2.84, RMSEA of 0.07 (90%CI 0.00-0.15), CFI of 0.99, TLI of 0.96, and SRMR of 0.02], acceptable internal consistency (Kuder-Richardson coefficient of 0.73 and McDonald’s omega of 0.74), excellent nomological validity (*r*_*s*_≥0.30 with SDS, SAS, CAT and FiND)] and non-Gender-DIF (t≤ 0.20).

In conclusion, the PSQ4D’s one-dimensional structure shows acceptable dimensionality, good reliability, acceptable nomological network, and non-Gender-DIF among Colombian COPD patients. Establishing the cutoff point with the best sensitivity and specificity is necessary.

## 1. Introduction

### 1.1 Depression

Depression is the most common mental disorder in the older population, with a negative impact on people’s quality of life and the economy of countries due to the lost years of healthy life (Cheng et al., 2024). The prevalence of depression is usually significantly higher in people who face acute and chronic stressors (Zenebe et al., 2021).

Therefore, the number of cases of depression is high in people with chronic diseases such as chronic obstructive pulmonary disease [COPD] (Atlantis et al., 2013; Matte et al., 2016). The examination for depressive symptoms is often omitted and, consequently, the diagnosis of depression among older COPD adults (Zareifopoulos et al., 2019). However, there are instruments for screening cases of depression with acceptable clinimetric performance (Lakkis & Mahmassani, 2015).

### 1.2 Primary care Screening Questionnaire for Depression (PSQ4D)

Indu et al. (2017) recently introduced the Primary care Screening Questionnaire for Depression (PSQ4D). The PSQ4D is a four-item instrument with a dichotomous response option, yes or no; the doctor can administer that and includes the key symptoms for the diagnosis of depression, depressed mood and anhedonia, and two psychosomatic symptoms of the disorder, such as feeling of fatigue and insomnia, associated symptoms included for the diagnosis of a major depressive episode in the International Classification of Diseases (World Health Organization, 2022) and the Diagnostic and Statistical Manual (American Psychiatric Association, 2022).

In India, the PSQ4D administered by primary care physicians to 827 primary care patients showed a sensitivity of 96%, specificity of 87%, positive predictive value of 74%, negative predictive value of 98%, and Cohen’s kappa of 0.72 at a score ≥ 2 compared to an interview conducted by a psychiatrist based on ICD-10 criteria. Moreover, the PSQ4D showed Cronbach’s alpha of 0.80 in the entire sample, with similar values according to age and gender (Indu et al., 2017). However, dimensionality, internal consistency, nomological network, and gender differential item functioning are unknown.

### 1.3 Practical issues

It is essential to know these properties of a brief instrument for depression screening because they are indirect indicators of its possible practical usefulness (Streiner et al., 2014), even more so in low—and middle-income countries in which there is little financial investment in a validation study using a clinical interview as a reference standard (Mari et al., 2006). Furthermore, it must be remembered that validating health measurement and screening instruments is a continuous and permanent process, given that widely divergent findings can be presented in different populations and over time (Streiner et al., 2014).

The study aimed to test the dimensionality, internal consistency, nomological network, and gender differential item functioning among elderly Colombian COPD patients.

## 2. Method

### 2.1 Design and participants

A psychometric study was conducted with adult COPD patients over 60 years who did not exhibit evident neurocognitive impairment or limitations in completing the self-administered questionnaires.

There was a non-probabilistic sample of 349 patients aged between 60 and 98 (M=75.56, SD=8.41). 61.89% were male participants, 79.37% had high school or less formal education, 88.57% were married or living in a free union, 73.92% dwelled in a low-income neighborhood, and 80.23% presented low deterioration (A or B COPD combined evaluation).

### 2.2 Measurements

#### 2.2.1 PSQ4D

The PSQ4D was applied as a self-report rating scale. It includes four items with a dichotomous response pattern that explore mood during the two most recent weeks (Spanish translation):

1. Have you been experiencing sadness or depressed mood, during the last two weeks or longer?

2. Have you been experiencing loss of interest or loss of pleasure in doing things, during the last two weeks or longer?

3. Have you been feeling excessively tired or without energy, during the last two weeks or longer?

4. Have you been suffering from sleeplessness during the last two weeks or longer?

Two or more affirmative answers suggest depression risk. The PSQ4D has shown acceptable clinimetric indicators: a specificity of 0.96 and a sensitivity of 0.87 (Indu et al., 2017).

#### 2.2.2 Brief Zung’s Self-rating Depression Scale (BSDS)

The BSDS is a ten-item tool that measures depressive symptoms during the most recent month. It offers four options, from “never” to “always,” that are rated from zero to three, depending on whether they suggest a major depressive episode. Possible scores are between zero and thirty; the higher the score, the greater the risk of depression (Campo et al., 2006). In the current sample, the scale showed acceptable internal consistency (Cronbach’s alpha of 0.72).

#### 2.2.3 Brief Zung’s Self-rating Anxiety Scale (BSAS)

The BSAS is a ten-point instrument that explores symptoms of anxiety over the past month. The scale presents four options from “never” to “always” that are rated zero to three, depending on whether they suggest an anxiety disorder. Total scores can be between zero and thirty; the higher the score, the greater the risk of anxiety (De La Ossa et al., 2009). Anxiety was included as one of the indicators of the nomological network for PSQ4D because anxiety scores are usually highly correlated with depression scores (Yohannes et al., 2022). This scale showed high internal consistency (Cronbach’s alpha of 0.82) in the present patient sample.

#### 2.2.4. COPD Assessment Test (CAT)

The CAT is an eight-item instrument to assess the impact of COPD on quality of life. Six response options are rated from zero to five, from lowest to highest severity. The total scores are between zero and forty; the higher the score, the more significant the deterioration in quality of life (Jones et al., 2009). This measurement was included for the nomological network because depression scores are usually related to quality-of-life scores or depressive symptoms, explaining a significant variance in the CAT score (Miravitlles et al., 2018). The CAT showed an acceptable internal consistency, Cronbach’s alpha of 0.73, in the current patients.

#### 2.2.5. Frail Non-Disabled [FiND] Screening Tool

The FiND is a questionnaire to measure frailty. It includes five items, the first two for ‘disability’ and the remaining three for ‘frailty.’ Each item has two scoring options: zero or one. The possible scores range between zero and five; the higher the score, the greater the risk of disability/fragility (Cesari et al., 2014). Frailty was included as one of the indicators of the nomothetic network for PSQ4D because the scores of that construct present a statistically significant correlation with the scores with different instruments that quantify the depression risk (Soysal et al., 2017). The FiND presented a Kuder-Richardson coefficient, equivalent to Cronbach, of 0.65 in the present study.

### 2.3 Procedure

It was conducted using a standard translation and back-translation process (International Test Commission, 2017). Two bilingual psychiatry professionals independently translated the PSQ4D from English to Spanish, and high agreement was observed. Minor discrepancies were resolved by consensus. Later, a translation professional translated the Spanish version into English. The comparison of the original English version and the back-translation product was highly concordant.

The patients were contacted in the specialized outpatient clinic of two Santa Marta, Colombia, institutions. The city has approximately 500,000 inhabitants and is located in the Colombian Caribbean, north of the country. On July 1, 2021, and June 30, 2022, patients completed the PQS4D in the office’s waiting room. After minimal instruction, this instrument can be completed in less than two minutes for people with five years of basic primary education.

### 2.4 Data analysis

#### 2.4.1 Dimensionality

Kaiser-Meyer-Olkin’s measure of sampling adequacy [KMO] (Kaiser, 1974), Bartlett’s test of sphericity (Bartlett, 1950), factor loadings, the eigenvalue, and the variance explained by the factor were calculated in the exploratory factor analysis.

In the confirmatory factor analysis, goodness-of-fit indicators were calculated: chi-square, normalized chi-square, root mean square error of approximation (RMSEA) and 90% confidence interval (90%CI), Comparative Fit Index (CFI), Tucker-Lewis Index (TLI) and standardized root mean square residual (SRMR). The chi-square probability was expected to be greater than 0.05, the normalized chi-square less than three (Hair et al., 2006), RMSEA less than 0.08 (Marcoulides & Yuan, 2017), SRMR less than 0.05, and CFI and TLI greater than 0.90 (Hu & Bentler, 1999).

#### 2.4.2 Internal consistency

Internal consistency was estimated using the Kuder-Richardson coefficient [K-RC] (Kuder & Richardson, 1937) and McDonald’s omega (McDonald, 1970). When the scale has a dichotomous response pattern, the K-RC is used instead of Cronbach’s alpha (Cronbach, 1951).

McDonald’s omega is a more reliable coefficient than Cronbach’s alpha if the tau equivalence principle is violated, which is necessary to accurately estimate Cronbach’s alpha (Campo-Arias & Oviedo, 2008).

#### 2.4.3 Nomological network

The PSQ4D total score was correlated with the total scores on BSDS, BSAS, CAT, and FiND using Pearson’s correlation coefficient (Pearson, 1909) or Spearman rank correlation (Spearman, 1910), depending on the symmetry of the distribution.

Besides, the mean and standard deviation of the PQS4D scores were compared according to gender and grade in the combined evaluation of COPD with the Student t-test (1908), after checking the homogeneity of variance with Levene’s F test. It was hypothesized that scores in women and patients with more severe COPD (C or D combined evaluation) would be higher than in men and less severely ill patients (A or B combined evaluation).

#### 2.4.4 Gender differential item functioning (Gender-DIF)

The Gender-DIF was estimated according to the tau-b correlation coefficient, indicated when one of the variables involved is dichotomous (Kendall, 1938). Correlations greater than 0.20 were considered the differential functioning acceptance criterion (Hambleton, 2006). The analysis was performed in the STATA.

## 3. Results

The PSQ4D scores were between zero and four (*M* = 1.07, *SD* = 1.32), and 109 patients (31.23%) were at risk for depression. The description of the performance of the items is presented in Table 1.

**Table 1.** Item description of the PQS4D.

| Item | *M* | *SD* | Kurtosis | Skewness | Communality | Loading |
| --- | --- | --- | --- | --- | --- | --- |
| Sadness | 0.32 | 0.47 | 0.76 | -1.44 | .62 | .79 |
| Loss of interest | 0.34 | 0.43 | 1.22 | -0.52 | .62 | .79 |
| Tired | 0.26 | 0.44 | 1.13 | -0.73 | .31 | .55 |
| Sleeplessness | 0.25 | 0.43 | 1.18 | -0.61 | .18 | .42 |

### 3.1. Dimensionality

The exploratory factor analysis showed a single salient dimension, with a KMO value of 0.73 and a Bartlett test with a chi-square of 317.17 (degrees of freedom of 6 and p-value of 0.001). The retained factor showed an Eigenvalue of 2.23, which explained 55.85% of the total variance.

Confirmatory factor analysis confirmed the one-dimensionality of the PSQ4D with acceptable goodness-of-fit indicators (Chi-square of 5.78 [degrees of freedom of 6 and p-value of 0.001], normalized chi-square of 2.84, RMSEA of 0.07 [90% CI 0.00 -0.15], CFI of 0.99, TLI of 0.96 and SRMR of 0.03).

### 3.2 Internal consistency

The PQS4D showed acceptable internal consistency with Cronbach’s alpha of 0.73 and McDonald’s omega of 0.74.

### 3.3 Nomological network

The nomological network was good. The PQS4D reported an appropriate correlation with SDS (*r*_*s*_=0.41), SAS (*r*_*s*_=0.43), CAT (*r*_*s*_=0.35), and FiND (*r*_*s*_=0.34).

Additionally, patients with more severe disease (C or D combined assessment showed higher scores than patients with less severe disease (A or B combined assessment) [M=1.43 (SD=1.43) versus 0.98 (SD=1.27), Levene’s F test=5.61, p-value =0.03, t=2.44, df=96.16, p-value=0.02]. However, the scores were similar in women and men [M=1.20 (SD=1.34) versus 0.98 (SD=1.30), Levene’s F test=2.01, p-value =0.16, t=1.53, df=347, p-value=0.13].

### 3.4 Gender-DIF

Finally, Gender-DIF analysis was not significant: Item 1 (sadness) showed t=0.10, item 2 (loss of interest) t=0.10, item 3 (tired) t=0.02, and item 4 (sleeplessness) t=0.03.

## 4. Discussion

The current report presents that the PSQ4D is a one-dimensional structure tool showing adequate dimensionality, good reliability, acceptable nomological network, and non-Gender-DIF among Colombian COPD patients.

This report is the first study to show that the PQS4D items converge on a single dimension. This finding is desirable for an instrument with only four items, given that a two-dimensional structure, two items for each dimension, could be irreplicable in future factor analysis (Streiner et al., 2014).

Likewise, the present study observed an internal consistency above 0.70, calculated with the Kuder-Richardson coefficient (0.73) and McDonald’s omega (0.74). Indu et al. (2017) documented a higher value of Cronbach’s alpha (0.80); However, Cronbach’s alpha is not the most appropriate coefficient to measure the internal consistency of a scale with a dichotomous response (Campo-Arias & Oviedo, 2008).

The nomological network and non-gender DIF found in this current study provide information on the construct validity and the possible practical usefulness of the PQS4D in patients with chronic diseases, such as COPD, in low- and middle-income countries, such as Colombia. These findings suggest that the instrument sufficiently approximates the diagnosis of a major depressive episode without gender bias. Biased symptom measurements are a critical problem in screening for mental health problems (Streiner et al., 2014).

### 4.1 Practical implications

Depression screening should be implemented in care services for patients with chronic conditions, such as COPD, who are at high risk of meeting the criteria for a major depressive episode (Atlantis et al., 2013; Matte et al., 2016). All screening must guarantee a timely and accurate diagnosis, effective treatment, and adequate follow-up (Whooley, 2016). These actions improve patients’ quality of life, reduce exacerbations and hospitalizations, and optimize the use of scarce resources allocated for mental health in most low- and middle-income countries (US Preventive Services Task Force et al., 2023).

### 4.2 Study’s strengths and limitations

This research has the strength of showing several indicators of the validity and reliability of the PQS4D in a sample of medically ill people. Likewise, the study shows the instrument’s performance in a version quickly adapted to Spanish due to the shortness of the questionnaire and the simplicity of the response to the items. Brevity and simplicity are relevant in context with people with limited reading and writing skills who can quickly become fatigued (Shrout & Yager, 1989).

Furthermore, the findings suggest that the instrument is equally valid and valuable as a self-administered scale. However, it has the limitation that the patients did not have a clinical interview with a psychiatrist to determine the best cutoff point in this population. The clinical interview is the validity criterion par excellence for clinical constructs such as depression (Streiner et al., 2014).

Another limitation is that similar scores were observed in men and women. However, readers should consider that the gender gap is narrowing in the prevalence of depressive symptoms in people over 60 years of age (Xu et al., 2024).This observation needs corroboration, given that scores on depression screening instruments usually show scores more suggestive of depression in women than men (Girgus et al., 2017; Zenebe et al., 2021).

### 4.3. Conclussions

The PSQ4D is a brief rating scale for screening depression with an acceptable one-dimensional structure, high internal consistency, satisfactory nomological network, and non-Gender-DIF among Colombian COPD patients. Further studies should establish the best cutoff point in this population.

## Data Availability

Data available on request from the authors.

